# High frequency and persistence of Anti-DSG2 antibodies in post COVID-19 serum samples

**DOI:** 10.1101/2022.02.23.22271045

**Authors:** Edward Lee, Ryan E. Tyler, Derrick Johnson, Natalie Koh, Ong Biauw Chi, Shi Yin Foo, Jack Tan

## Abstract

**Background:** There is growing recognition that COVID-19 does cause cardiac sequelae. The underlying mechanisms involved are still poorly understood to date. Viral infections, including COVID-19, have been hypothesized to contribute to autoimmunity, by exposing previously hidden cryptic epitopes on damaged cells to an activated immune system Given the high incidence of cardiac involvement seen in COVID-19, our aim was to determine the frequency of anti-DSG2 antibodies in a population of post COVID-19 patients.

**Methods and Results:** 300 convalescent serum samples were obtained from a group of post COVID-19 infected patients from October 2020 to February 2021. 154 samples were drawn 6 months post-COVID-19 infection and 146 samples were drawn 9 months post COVID infection. 17 samples were obtained from the same patient at the 6- and 9-month mark. An electrochemiluminescent-based immunoassay utilizing the extracellular domain of DSG2 for antibody capture was used. The mean signal intensity of anti-DSG2 antibodies in the post COVID-19 samples was significantly higher than that of a healthy control population (19+/−83.2 vs. 2.1+/−6.8, P value <0.001). Of note, 29.3% of the post COVID-19 infection samples demonstrated a signal higher than the 90th percentile of the control population and 8.7% were higher than the median found in ARVC patients. The signal intensity between the 6-month and 9-month samples did not differ significantly.

**Conclusions:** We report for the first time that recovered COVID-19 patients demonstrate significantly higher and sustained levels of anti-DSG2 autoantibodies as compared to a healthy control population, comparable to that of a diagnosed ARVC group.

---

The SARS-CoV-2 coronavirus has infected at least 300 million people worldwide, with ~5 million deaths to date attributable to the resulting disease COVID-19. There is growing recognition that COVID-19 may result in a variety of long-term sequelae, of which cardiac compromise may be the most under-recognized as its symptoms may be attributed to other organ systems.

COVID-19 has been associated with MRI evidence of myocardial involvement and arrhythmias well into recovery, independent of preexisting conditions, severity and overall course of the acute illness, and the time from the original diagnosis. The percentage of patients who develop a depressed ejection fraction after COVID-19 recovery are not well-understood, although frank cardiomyopathy has been described in post-COVID-19 patients. A recent study (PROLUN study; ref. 1) demonstrated right ventricular and diastolic dysfunction in approximately half of the patients, with arrhythmias in ~27%, 3 months after COVID-19 (1).

The findings of right-sided cardiomyopathy and increased predilection for arrhythmias are also features of arrhythmogenic right ventricular cardiomyopathy (ARVC). Antibodies to the desmosome protein desmoglein-2 (DSG2) have been shown to be present in some patients with ARVC (2). Concentrations of anti-DSG2 antibodies correlate positively to arrhythmia burden, and presence of these antibodies in borderline ARVC cases predicts the development of fulminant ARVC (2). Exposure of cardiomyocytes to anti-DSG2 antibodies in vitro results in a reduction in gap junction function. Together, these data suggest that anti-DSG2 antibodies may play a functional role in cardiac pathology.

Viral infections, including COVID-19, have been hypothesized to contribute to autoimmunity, e.g., by exposing previously hidden cryptic epitopes on damaged cells to an activated immune system (3). Given the high incidence of cardiac involvement seen in COVID-19, we hypothesized that anti-DSG2 autoantibodies might be generated as a result. We developed an electrochemiluminescent-based immunoassay utilizing the extracellular domain of DSG2 for antibody capture; assay performance was validated by appropriate capture of commercially available anti-DSG2 antibodies (polyclonal goat anti-human DSG2 antibody; R&D Systems) (Fig. 1A).

**Figure 1.**
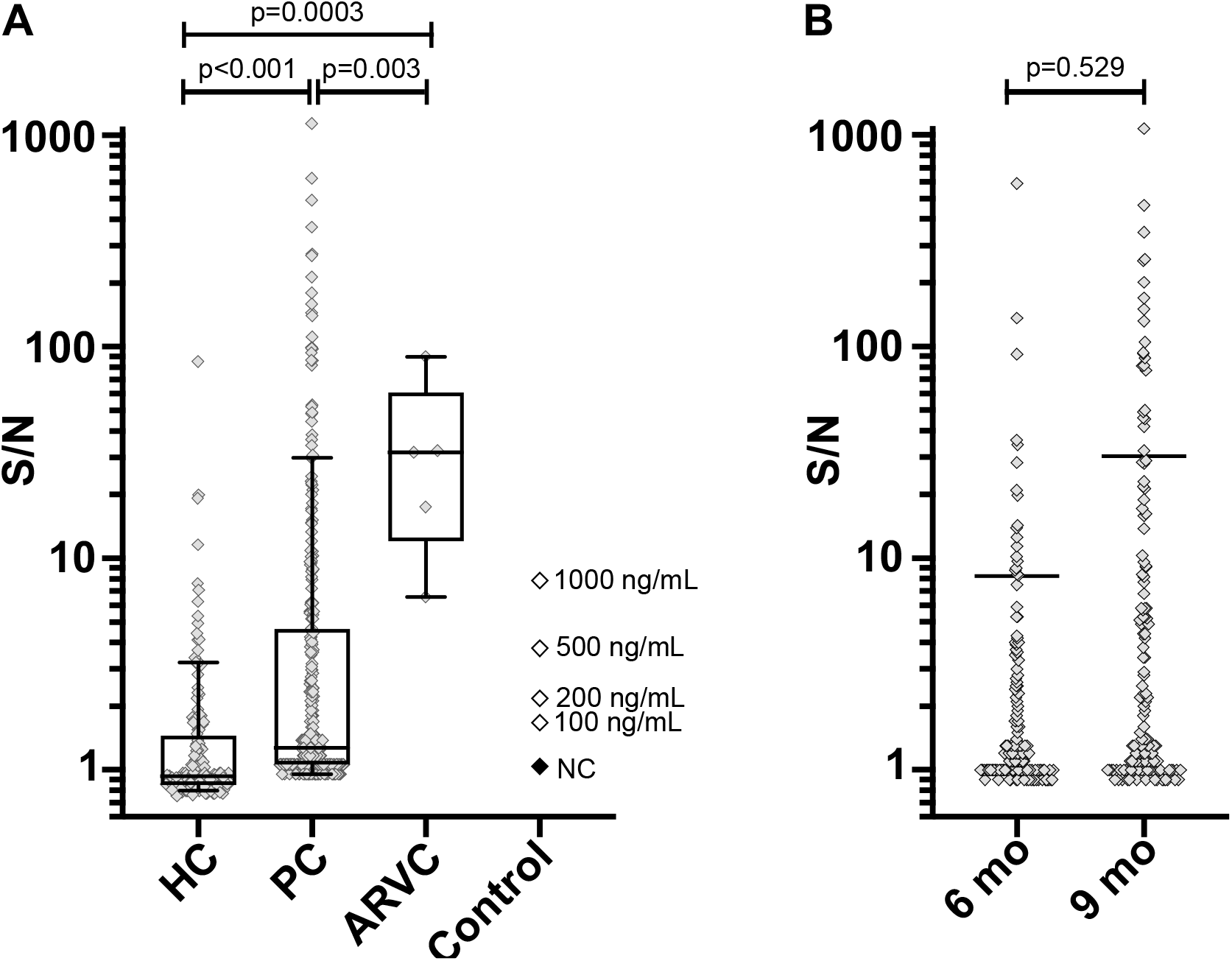
**A**. Comparative levels of anti-DSG2 antibody signal in healthy controls (N=152), post-COVID-19 (N=300) and arrhythmogenic right ventricular cardiomyopathy samples (N=5). **B**. Comparison of levels of anti-DSG2 antibody signals in the PC (total N=300) group by 6 (N=154) and 9 (N=146) months after COVID-19 infection. HC, healthy controls; PC, post-COVID-19; ARVC, arrhythmogenic right ventricular cardiomyopathy; Control, positive control goat anti-DSG2 polyclonal antibody (R&D Systems, AF947) or NC, negative control pool of non-reactive human serum; S/NC, signal/negative control; individual grey diamonds represent a single serum sample each; box and whisker limits represent 25^th^-75^th^ and 10^th^-90^th^ percentiles, respectively. Average assay values for 100, 200, 500 and 1,000 ng/mL of control anti-DSG2 polyclonal antibody are designated with white diamonds. The assay value for the negative control is designated with a black diamond. B. Comparative levels of anti-DSG2 antibody in 6 months (n=154) and 9 months (n=146) post-COVID-19 samples. 6 mo, 6 months post-COVID; 9 mo, 9 months post-COVID. Average signal intensities are designated with black bars. P-values based on non-parametric rank-based Wilcoxon-Mann-Whitney 2-sided test.

300 convalescent serum samples were obtained from a group of post-COVID-19 infected patients from October 2020 to February 2021 as part of an ongoing epidemiological study of a young, East Asian-populated dormitory in Singapore. The mean age of our study population was 37 years old (range 21-65). 154 samples were drawn 6 months post-COVID-19 and 146 samples were drawn 9 months post COVID. 17 samples were obtained from the same patient at the 6- and 9-month mark. The negative control group sera were obtained from a commercial source (BioIVT) of self-declared healthy individuals. Positive control ARVC sera were obtained from a prior study (4). The mean signal intensity of anti-DSG2 antibodies in the post COVID-19 samples was significantly higher than that of a healthy control population (19**±**83.2 in the post COVID-19 sample vs. 2.1**±**6.8 in the healthy control population, P value <0.001). Of note, 29.3% of the post COVID-19 samples demonstrated a signal higher than the 90^th^ percentile of the control population and 8.7% were higher than the median found in ARVC patients. The signal intensity between the 6-month and 9-month samples did not differ significantly (p=0.529; Fig. 1B). The caveat to this comparison is that the separate groups of samples (post-COVID-19, healthy controls and ARVC sera) were assessed non-contemporaneously; however, same-group repeat testing (for positive controls, healthy controls and ARVC groups) has demonstrated high repeatability; negative control samples demonstrated minimal variation across tests and were used to normalize all data.

We report for the first time that recovered COVID-19 patients demonstrate significantly higher levels of anti-DSG2 autoantibodies, and that these antibody levels are sustained well into recovery from COVID-19 – up to 6 and 9 months. While other groups (4) have demonstrated increased overall levels of autoantibodies during the acute/subacute phases of COVID-19, our data demonstrates the prolonged and robust elevation of a specific autoantibody; in addition, as anti-DSG2 antibodies from ARVC patients appear to cause direct cardiac pathology in vitro (2), this has implications for long-term, post-COVID-19 cardiac compromise. Of note, ~29% of our patients had levels of anti-DSG2 autoantibodies above the 90^th^ percentile of a comparator normal control group, whereas ~27% of post-COVID-19 patients had notable arrhythmia in the PROLUN stud (1).

The limitations to this study are that we were unable to analyze the relationship between anti-DSG2 autoantibody levels and current symptoms, given the lack of available clinical data for these patients, and that it is not known if COVID-19 vaccinations generate similar frequencies of anti-DSG2 antibodies. The presence of anti-DSG2 antibodies after COVID-19 recovery may have important risk stratification implications in determining vocational suitability and fitness for competitive sports. Further work is required to demonstrate that the anti-DSG2 autoantibodies found in post-COVID-19 patients have direct cardiotoxicity.

## Data Availability

All data produced in the present work are contained in the manuscript

